# Structural racism and COVID-19 response: Higher risk of exposure drives disparate COVID-19 deaths among Black and Hispanic/Latinx residents of Illinois, USA

**DOI:** 10.1101/2021.08.04.21261595

**Authors:** Tobias M Holden, Melissa A. Simon, Damon T. Arnold, Veronica Halloway, Jaline Gerardin

## Abstract

**BACKGROUND:** Structural racism has driven and continues to drive policies that create the social, economic, and community factors resulting in residential segregation, lack of access to adequate healthcare, and lack of employment opportunities that would allow economic mobility. This results in overall poorer population health for minoritized people. In 2020, Black and Hispanic/Latinx communities throughout the United States, including the state of Illinois, experienced disproportionately high rates of COVID-19 cases and deaths. Public health officials in Illinois implemented targeted programs at state and local levels to increase intervention access and reduce disparities.

**METHODS:** To quantify how disparities in COVID outcomes evolved through the epidemic, data on SARS-CoV-2 diagnostic tests, COVID-19 cases, and COVID-19 deaths were obtained from the Illinois National Electronic Disease Surveillance System for the period from March 1 to December 31, 2020. Relative risks of COVID-19 cases and deaths were calculated for Black and Hispanic/Latinx vs. White residents, stratified by age group and epidemic interval. Deaths attributable to racial/ethnic disparities in incidence and case fatality were estimated with counterfactual simulations.

**RESULTS:** Disparities in case and death rates became less drastic after May 2020, but did not disappear, and were more pronounced at younger ages. From March to May of 2020, the risk of a COVID-19 case for Black and Hispanic/Latinx populations was more than twice that of Whites across all age groups. The relative risk of COVID-19 death reached above 10 for Black and Hispanic/Latinx individuals under 50 years of age compared to age-matched Whites in the early epidemic. In all Illinois counties, relative risk of a COVID-19 case was the same or significantly increased for minoritized populations compared to the White population. 79.3% and 86.7% of disparities in deaths among Black and Hispanic/Latinx populations, respectively, were attributable to differences in age-adjusted incidence compared to White populations rather than differences in case fatality ratios.

**CONCLUSIONS:** Racial and ethnic disparities in the COVID-19 pandemic are products of society, not biology. Considering age and geography in addition to race/ethnicity can help to identify the structural factors driving poorer outcomes for certain groups. Studies and policies aimed at reducing inequalities in disease exposure will reduce disparities in mortality more than those focused on drivers of case fatality.

## Introduction

By the end of 2020, the United States (US) had reported more cases of SARS-CoV-2 infection and COVID-19 deaths than any other country [1]. Minoritized groups, particularly Black and Hispanic/Latinx communities, suffered an outsized share of infection and mortality burden in the US [2]. Structural racism includes decades of disinvestment and racist policies in multiple sectors, which further socioeconomic and health inequality in minoritized populations [3]–[5]. The resultant social determinants serve as drivers of health disparities that are particularly salient in infectious disease outbreaks. For a respiratory virus like SARS-CoV-2, structural racism drives systemic differences in both exposure via working and living conditions, and case fatality via prevalence of underlying conditions that increase the likelihood of severe disease [6].

Illinois is the fifth most populous US state with Chicago being the third most populous city in the US. Demographically, the state’s age distribution matches that of the wider US, and the percent of the population identifying as Hispanic, non-Hispanic Black/White/Asian/Native American/Other, or more than one race are each within 2% of national estimates [7]. Though diverse, Illinois has high indices of residential segregation between counties and within metropolitan areas [7], perhaps one of the most obvious manifestations of structural racism. A study of mobility patterns in Chicago and other major US cities from March to May 2020 found that people from lower-income and non-white census block groups may have higher infection rates because they had not been able to reduce mobility to the same degree, and tended to visit more crowded locales when mobile [8].

Structural factors beyond residential segregation predispose minoritized populations to COVID-19 exposure, and the influence of each likely varies across neighborhoods, age groups, and throughout the course of an epidemic. Following significant community transmission of SARS-CoV-2 and hospitalizations due to COVID-19, the governor of Illinois issued a state-wide “Stay-At-Home” order beginning March 21, 2020, which deemed certain occupations “essential work”, and therefore exempt from the requirement to stay home [9]. In the Chicago Metropolitan area, 54.1% of essential workers are people of color, compared to 43.9% of all workers [10]. Relative to their share of the civilian workforce, Black workers make up a larger share of firefighters, community/social service workers, and transportation, healthcare support, and material moving occupations state-wide. Hispanic/Latinx workers are overrepresented in production, material moving, construction and extraction, cleaning and maintenance, and food preparation and food service occupations [11]. Several outbreaks in Illinois have been tied to food preparation and meat processing plants [12]. Another early study linked jail cycling in Cook County, where roughly 75% of inmates are Black and 16% are Hispanic, with 15% of all COVID-19 cases in the state of Illinois [13].

Analysis of disparities that accounts for age and geography can be useful for assessing their structural causes and developing focused interventions to promote justice in public health. In response to early data on racial and ethnic disparities in COVID-19 cases and deaths, public health officials were able to implement directed community programs, improving access to key interventions such as diagnostic testing. In this study, we analyze COVID-19 testing, case, and death data from Illinois across three epidemic intervals from March 1 to December 31, 2020, by race/ethnicity, age, and county to observe the evolution of disparities amidst changing surveillance strategies and public health policies.

## Methods

### Datasets

Population counts, including county population by age and race/ethnicity, were obtained from the 2018 American Community Survey (2018 ACS) [14].

Confirmed COVID-19 case data (“case data”) were obtained from the Illinois National Electronic Disease Surveillance System (I-NEDSS) database and included all individuals with diagnosed SARS-CoV-2 infection reported to the Illinois Department of Public Health (IDPH). The data for each case included home ZIP code, county, sex, race, ethnicity, dates of specimen collection, hospital admission and death (where applicable), and whether a patient died from COVID-19. Deaths were attributed to COVID-19 in I-NEDSS based on any of the following: 1) the presence of COVID-19 on a death certificate; 2) death within 30 days of diagnosis/symptom onset or during hospitalization, unless cause of death is clearly unrelated to COVID-19, such as homicide or accident; 3) diagnosis of COVID-19 without return to baseline health; or 4) an autopsy result consistent with COVID-19.

We began with an initial sample of all COVID-19 cases recorded in I-NEDSS and received from IDPH on February 10, 2021 (Table 1), which we then restricted to those with specimen collection dates between March 1, 2020, and December 31, 2020. We defined admissions as the subset of cases with hospital admission indicated in I-NEDSS or with a valid date of admission before February 10, 2021. We defined COVID-19 deaths as the subset of these cases with a recorded date of death before February 10, 2021, and attribution of death to COVID-19. We estimate this 41-day lag to encompass 95% of confirmed COVID-19 deaths, based on the distribution of time from first positive specimen to deceased date in this dataset.

**Table 1.**
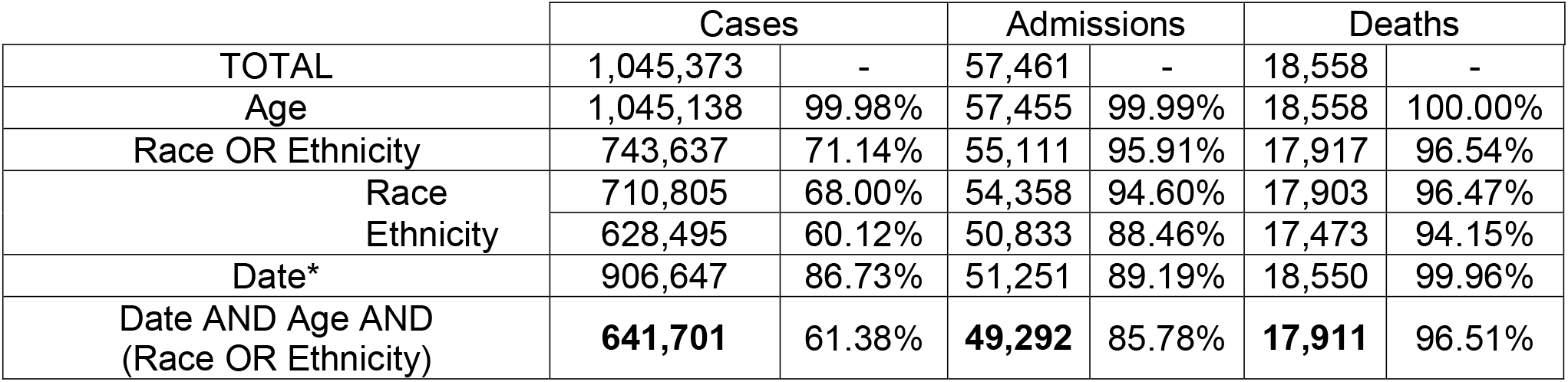
Description of individual COVID-19 case data sample obtained from I-NEDSS on February 10, 2021, including all cases with specimen collection date between March 1 and December 31, 2020. *”Date” row cutoff – specimen collection: Dec 31, 2020, admission/death: Feb 10, 2021.

|  | Cases |  | Admissions |  | Deaths |  |
| --- | --- | --- | --- | --- | --- | --- |
| TOTAL | 1,045,373 | - | 57,461 | - | 18,558 | - |
| Age | 1,045,138 | 99.98% | 57,455 | 99.99% | 18,558 | 100.00% |
| Race OR Ethnicity | 743,637 | 71.14% | 55,111 | 95.91% | 17,917 | 96.54% |
| Race | 710,805 | 68.00% | 54,358 | 94.60% | 17,903 | 96.47% |
| Ethnicity | 628,495 | 60.12% | 50,833 | 88.46% | 17,473 | 94.15% |
| Date* | 906,647 | 86.73% | 51,251 | 89.19% | 18,550 | 99.96% |
| Date AND Age AND<br>(Race OR Ethnicity) | **641,701** | 61.38% | **49,292** | 85.78% | **17,911** | 96.51% |

Race and ethnicity are reported as separate variables in I-NEDSS. To account for this while avoiding small sample sizes, we added a new racial category of “Hispanic/Latinx” encompassing all cases with Hispanic/Latinx ethnicity, regardless of race. All other cases were considered non-Hispanic/Latinx and categorized according to the race variable given in I-NEDSS. Cases indicated as belonging to multiple racial groups were categorized as “Other”, unless Hispanic/Latinx ethnicity was also indicated. Cases with race reported as “Unknown” or not reported and ethnicity reported as “Unknown” or “Not Hispanic/Latinx” were categorized as “Unknown”. To evaluate the impact of assuming all cases of unknown ethnicity were non-Hispanic/Latinx, a sensitivity analysis was performed (see supplement) in which cases with unknown ethnicity were either excluded or assumed to be Hispanic/Latinx.

COVID-like illness (CLI) admissions data were received from IDPH on February 10, 2021.

Data on SARS-CoV-2 diagnostic testing in Illinois were received from IDPH (“test data”) on February 10, 2021 (Table 2). Diagnostic tests included PCR and antigen tests but not serological tests. The test data was aggregated as daily total tests and positive tests by age and race/ethnicity. In this dataset, race and ethnicity were already recorded as a single variable, including Hispanic/Latinx as one group. We restricted our demographic analysis to diagnostic testing among people ages 0-100. Multiple tests on the same individual were counted equally, so positive tests in the test data do not directly correspond to cases in the case data.

**Table 2.** Description of SARS-CoV-2 diagnostic testing data obtained from I-NEDSS on February 10, 2021, including total tests administered and positive test results between March 3 and December 31, 2020.

|  | Total Tests |  | Positive Tests |  |
| --- | --- | --- | --- | --- |
| TOTAL | 12,746,916 | - | 1,131,284 | - |
| Age | 12,671,596 | 99.41% | 1,125,331 | 99.47% |
| Race/Ethnicity | 7,143,080 | 56.04% | 652,643 | 57.69% |
| Age AND Race/Ethnicity | **7,109,855** | 55.78% | **649,493** | 57.41% |

All moving averages were 7-day lagged moving averages.

### Relative risks

Cases were divided into 3 intervals based on specimen collection date: March 1 to May 31 (initial outbreak and control), June 1 to September 30 (lower-incidence summer), and October 1 to December 31, 2020 (fall resurgence). Relative risks were calculated as the ratio between the cumulative per capita case or death rates for Hispanic/Latinx or Black individuals and age-matched White individuals who tested positive during the same interval.

### Mapping COVID-19 case disparities by county

Case data were paired with county demographic data by matching patient county of residence to counties in the 2019 American Community Survey (ACS). All cases with known race/ethnicity and a county within Illinois were included. 892 cases were excluded from county-level analysis due to the county of residence being missing or outside Illinois. An additional 264,556 cases of “Unknown” race/ethnicity in valid counties were also excluded.

We estimated disparities in each county using the ratio between per capita case rates for age-matched White and non-White residents (naive relative risk) during each epidemic interval. Relative risk was not calculated for counties with fewer than 10 White and 10 non-White cases in any given age group and epidemic interval. 95% confidence intervals were used to determine significance.

### Counterfactual death estimation by matching incidence and case fatality ratios

Incidence rates were defined as COVID-19 cases per person per week. Case fatality ratios (CFR) were defined as the ratio of COVID-19 deaths to COVID cases, where cases were defined as all SARS-CoV-2 infections recorded in Illinois surveillance. This case definition excludes infections that were never diagnosed or reported. Deaths used in the numerator for CFR were all eventual deaths among the cases discovered that week. Weekly incidence, CFR, and mortality rates were smoothed with a 3-week centered rolling average. We simulated counterfactual scenarios in which minoritized populations were subject to the same weekly parameters of incidence, CFR, or mortality rate as age-matched White populations. Counterfactual weekly deaths were predicted in each scenario as

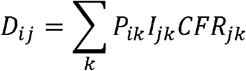

where *D*_*ij*_ is the weekly deaths in minoritized population *i* under scenario *j, P*_*ik*_ is the population of group *i* in age group *k, I*_*j*_ is the incidence in age group *k* in group *i* for the matched CFR scenario and in the White population for other scenarios, and *CFR*_*j*_ is the CFR in age group *k* in group *i* for the matched incidence scenario and in the White population for other scenarios.

Percent reduction in deaths was calculated between the deaths predicted in each scenario, and the total deaths under the curve generated from the smoothed weekly death rates.

## Results

### Racial and ethnic composition of COVID-19 cases, admissions, and deaths in Illinois

From March 1, 2020, to December 31, 2020, Illinois identified 906,647 cases of SARS-CoV-2 infection among its residents, 18,558 of whom died from COVID-19 by February 10, 2021 (Table 1).

Compared to their share of the population, individuals of Hispanic/Latinx, Native American, or “Other” race/ethnicity were overrepresented among Illinois COVID-19 cases (Figure 1A). Hispanic/Latinx, Black, and Native American populations were hospitalized and died at higher rates than their portion of overall state demographics, and Black people were overrepresented among admitted cases relative to their share of all cases. The combination of a lower share of cases but higher share of hospitalizations/deaths among Black people suggests either increased susceptibility to severe disease, under-testing, or both, in this population.

**Figure 1.**
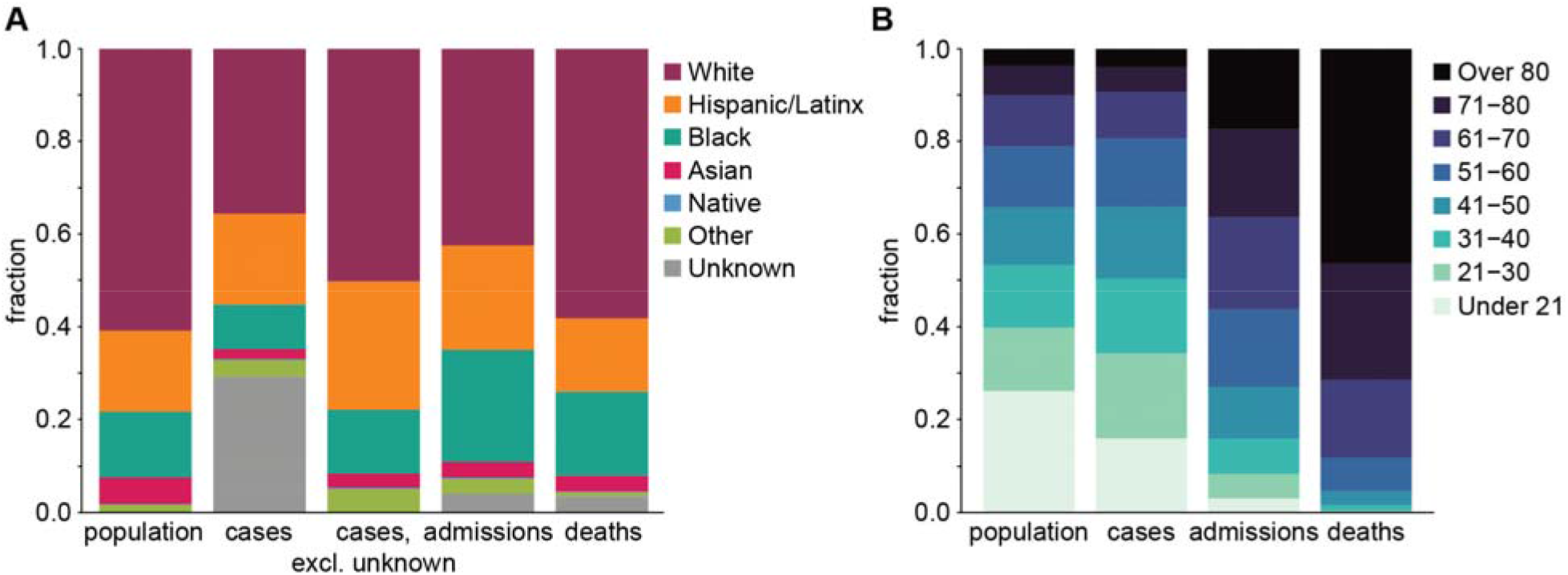
Demographics of cumulative COVID-19 cases, hospital admissions, and deaths in the state of Illinois through December 31, 2020, by A) race and ethnicity and B) age group, compared with the composition of the general population.

In Illinois, older adults and the elderly comprise a disproportionate share of admitted cases and deaths, while cases are only slightly skewed toward older individuals (Figure 1B). The overrepresentation of older adults among hospitalizations and deaths reflects the known association between age and severe COVID-19. More than 40% of all confirmed COVID-19 deaths among cases detected in this period have been traced to clusters in skilled nursing, rehabilitation, and long-term care facilities. Because the Hispanic/Latinx population is younger in Illinois than the Black population, which is younger than the White population (Figure S1), age effects should be accounted for when quantifying disparities in COVID-19 burden across racial and ethnic groups. Elderly adults over 70 years old are more than 75% White, while younger people under 40 are less than 60% White. Based on these patterns alone, we expect to see the proportion of admissions and deaths that are White to be greater than the White proportion of cases.

The COVID-19 epidemic in Illinois through December 31, 2020, can be divided into three intervals: the initial outbreak and containment from March through the end of May, when mitigations began to be lifted [15] (interval 1); a second period of lower transmission from June to October (interval 2); and a third surge in cases beginning in October (interval 3) (Figure 2). Interval 1 disproportionately impacted Black and Hispanic/Latinx residents, who comprised roughly half of all cases, hospitalizations, and deaths from March to June. Throughout the epidemic, White individuals comprise a larger share of hospitalizations and deaths than of cases, suggesting a consistently higher fraction of severe disease among White cases. The elevated case fatality and case hospitalization ratios in the White population is consistent with their older age structure but could also be due to under-detection of milder COVID-19 cases in the younger white population.

**Figure 2.**
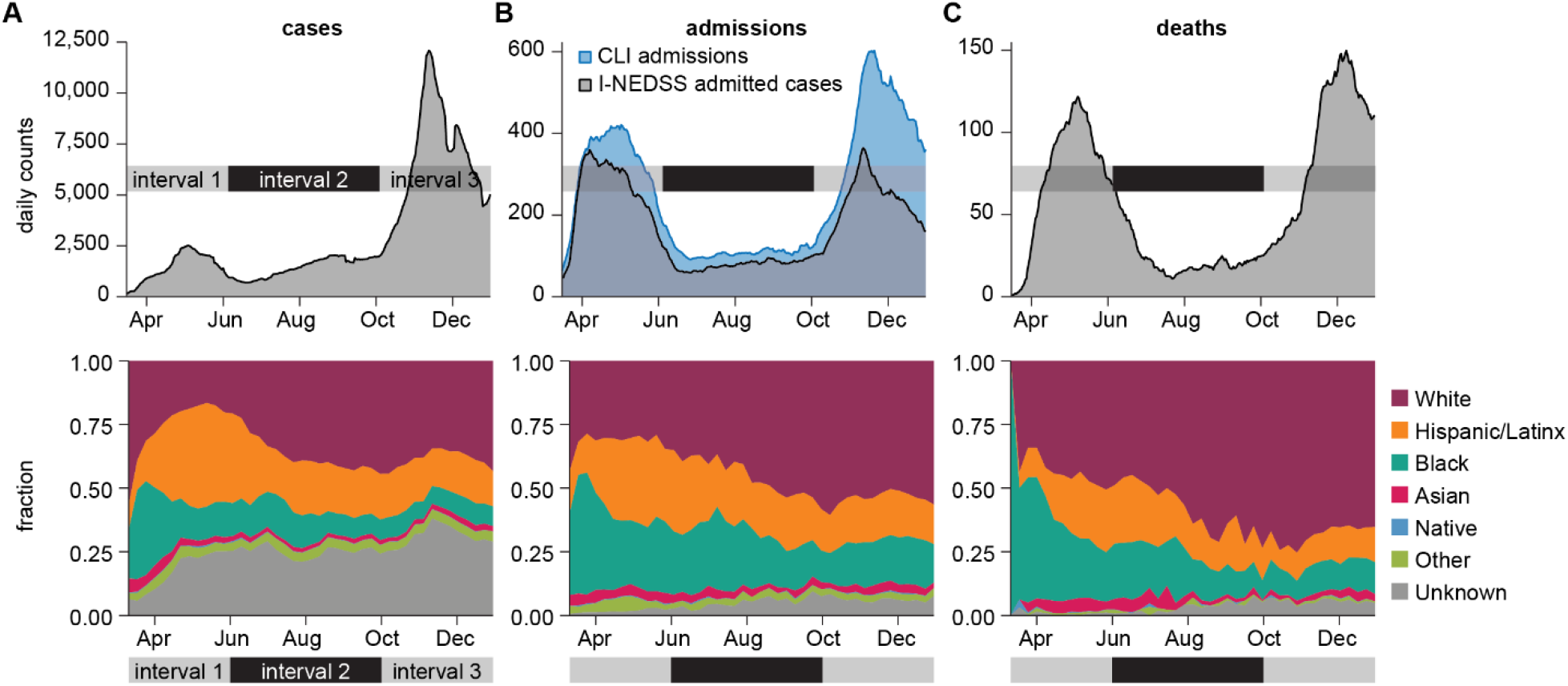
Changing racial and ethnic composition of COVID-19 cases, admissions, and deaths through three epidemic intervals. 7-day moving averages (top) and weekly fraction by race/ethnicity (bottom) are shown for each indicator from March 1 to December 31, 2020: A) cases B) admissions and C) deaths. COVID-like illness (CLI) admissions outnumber confirmed COVID-19 admissions in I-NEDSS.

Data quality limits the scope of analysis on racial and ethnic disparities in COVID-19. Race and ethnicity were not available for 29.2% of all cases, 4.1% of hospital admissions, and 3.5% of confirmed deaths. Since the peak of interval 1 in late April, the weekly portion of cases without reported race/ethnicity has ranged from 25% to 45%. Compared to COVID-like illness (CLI) admission data from hospitals, admitted cases from I-NEDSS are much lower during interval 3 and also lower during interval 1, suggesting limited completeness of this data field. The remaining analyses therefore focus on cases and deaths rather than admissions.

### Testing intensity by age and race/ethnicity

Illinois recorded 12,746,916 total tests and 1,131,284 positive results between March 3 and December 31, 2020, however race/ethnicity was not reported for more than 40% of the dataset (Table 2).

Reporting of COVID-19 cases relies on access to diagnostic testing, and differences in testing rates may contribute to differences in case counts. Testing was low for all groups during interval 1, especially prior to May, just exceeding 1 per 1,000 per day in the elderly White population and among Black and Hispanic/Latinx adults (Figure 3A). Test positivity rates (TPR) for Black and Hispanic/Latinx populations were much higher than in the White population at this time (Figure 3B), suggesting relatively insufficient testing in Black and Hispanic/Latinx demographics despite their slightly higher rate of per capita testing [16]. TPR exceeded 35% in all Hispanic/Latinx populations over 11 and Black populations over 18 in interval 1, compared with peak values of around 20% in White.

**Figure 3.**
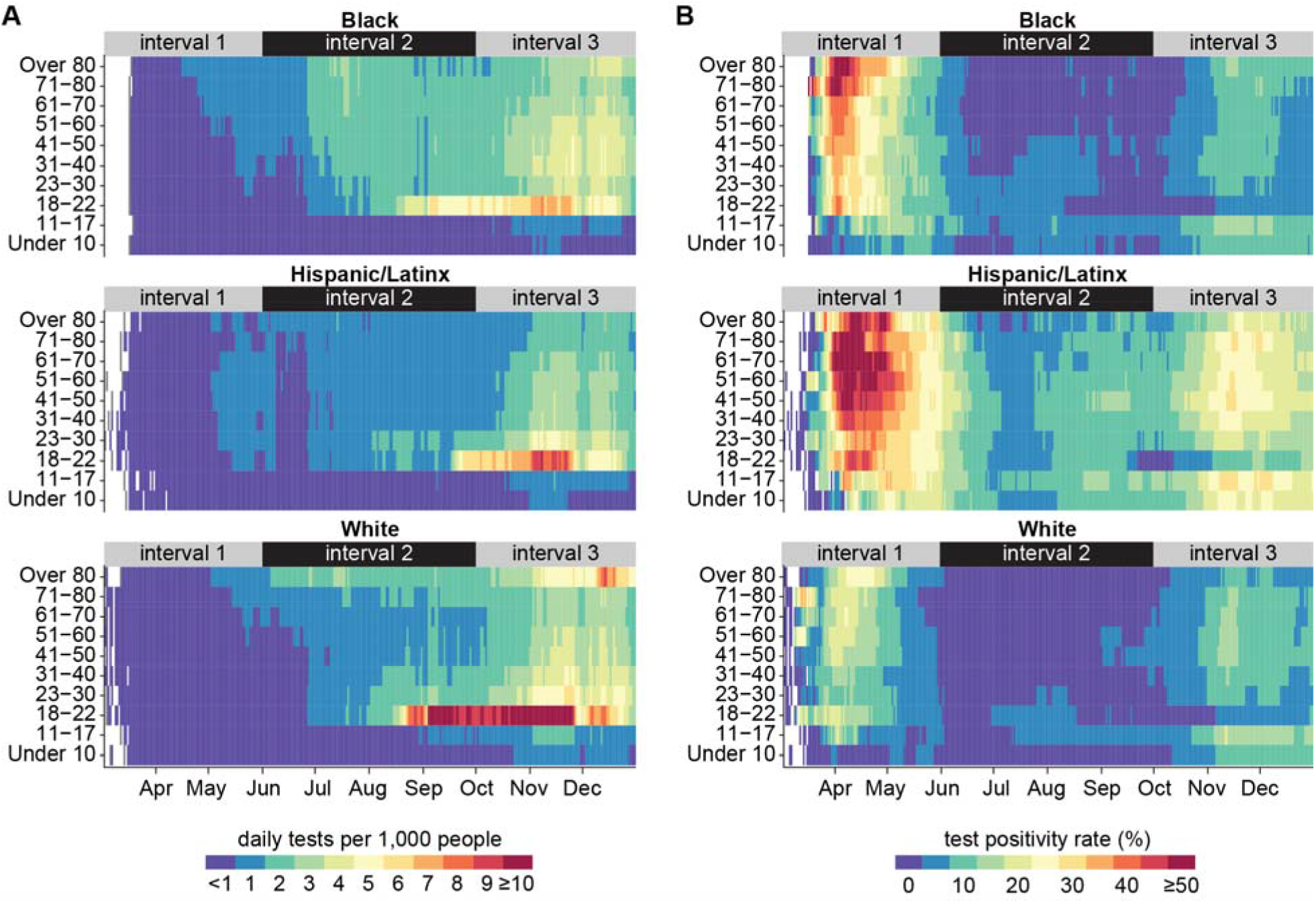
Diagnostic testing intensity and test positivity rate by age and race/ethnicity through December 31, 2020. A) 7-day moving average of tests administered per 1,000 population. B) 7-day moving average of test positivity rate.

In May, testing began to expand among Hispanic/Latinx people over 17 years of age, Black people over 22 years, and White people over 60 years, before dropping again for Black people under 41 and Hispanic/Latinx people Under 81 in June. In early July, testing increased sharply for Black adults, who had the highest per capita testing rates outside of the White people over 80. Testing rates for Hispanic/Latinx individuals did not increase as much despite this demographic experiencing the highest caseload during interval 1 and continued high TPR thereafter. During interval 2, test positivity rates remained around 5-10% for Hispanic/Latinx populations, indicating continuous community transmission of SARS-CoV-2.

Testing rates rose rapidly for people ages 18-22 beginning in August, coinciding with the return of students to college campuses where regular routine testing was implemented. The University of Illinois at Urbana-Champaign, for example, was testing its entire on-campus population of more than 25,000 people twice per week [17]. Daily testing rates increased to above 10 per 1,000 in for White people ages 18-22 and plateaued at 6 per 1,000 and 8 per 1,000 for Black and Hispanic/Latinx people ages 18-22 respectively. The higher testing per capita in this age group since September is reflected in their having the lowest test positivity rate (Figure 3B).

During interval 3, increases in per capita testing were accompanied by increases in TPR for all demographics, indicating that growth in testing failed to keep pace with the growing epidemic. In November, daily testing rates rose above 3 in 1,000 for adults in all three major racial and ethnic groups except Hispanic/Latinx people over 70. Testing remained lowest in the Hispanic/Latinx population, which was the only racial/ethnic group to see TPR rise above 20% and for some ages, even above 30% during interval 3.

### Temporal trends in age-adjusted disparities in risk of a COVID-19 case

The distribution of COVID-19 cases by age was not the same for the three largest racial and ethnic groups in Illinois. Smoothed daily incidence by race/ethnicity and age group is shown in Figure 4A. Incidence was highest for Hispanic/Latinx and Black people over 80, exceeding 7-8 per 10,000 per day between April and June, while rates remained below 3 in 10,000 for White people the same age. Meanwhile, daily incidence remained below 1 per 10,000 for White people under 80 years old, lower than for the rate for Black people over 20 and Hispanic-Latinx people in any age group. Incidence in interval 1 was skewed toward the elderly in the White population but less so for Black populations and much less so for Hispanic/Latinx populations.

**Figure 4.**
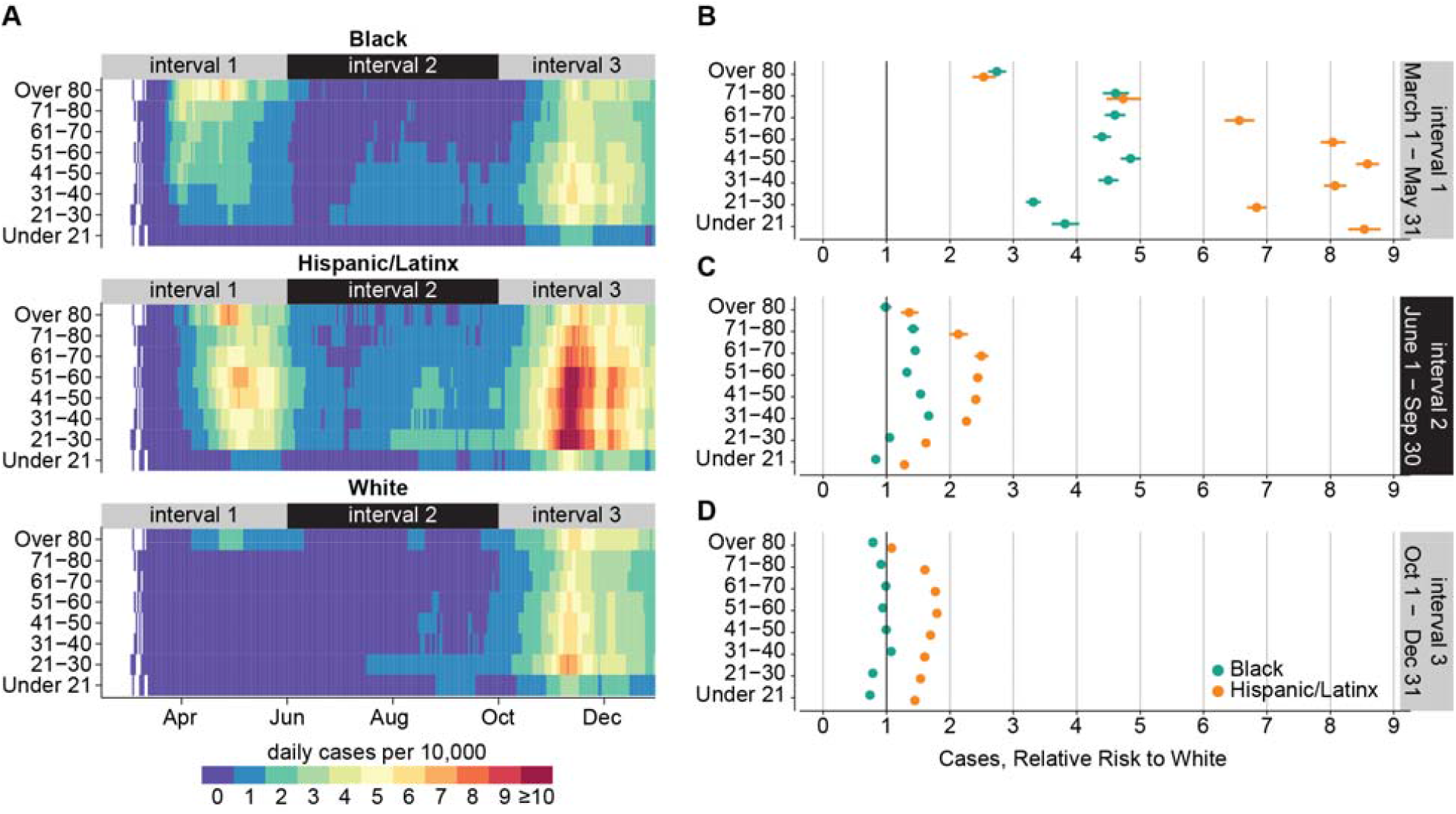
Age-adjusted disparities in risk of COVID-19 case by race/ethnicity from March 1 to December 31, 2020. A) 7-day moving average of COVID-19 cases per 10,000 population, by age and race/ethnicity. B-D) Relative risk of COVID-19 case over three epidemic intervals. Horizontal lines indicate 95% confidence intervals.

During interval 1, the risk of a COVID-19 case was 2.5-5x greater for Black and 2.5-9x greater for Hispanic/Latinx people compared to White people in the same age group (Figure 4B). Relative risks were lowest in the elderly and increased for younger age groups. For all age groups below 70, relative risks were higher for the Hispanic/Latinx population compared with White than for the Black population compared with White.

Compared with interval 1, incidence was lower in interval 2 (Figure 4A). Cases in interval 2 in the White population were nearly absent and limited to young adults, especially after universities began to reopen in August. Hispanic/Latinx individuals continued to experience the highest case burden, with incidence above 1 per 10,000 per day for all age groups and above 2 per 100,000 adults ages 21-60, from mid-July onward. In Black demographics, incidence was lower for older age groups, a reversal of the trend seen in the first interval.

Risk of a COVID-19 case in Black and Hispanic/Latinx populations relative to White also decreased in interval 2, although disparities persisted. Hispanic/Latinx individuals of all ages and Black individuals ages 31-80 were 1.5-2.5x more likely to experience a case of COVID-19 than age-matched White individuals during interval 2 (Figure 4C).

During interval 3, cases increased rapidly beginning in October and quickly exceeded peak numbers from interval 1 for most demographic groups. Compared with interval 1, cases in interval 3 were much younger. Daily incidence rose above 4 per 10,000 for all demographics older than 21. Black and White daily incidence was highest among adults under 50, rising above 5 and 6 per 10,000 respectively. Case burden was especially heavy in the Hispanic/Latinx population, with daily incidence above 10 per 10,000 for working-age adults (ages 21-60). The relative risk of a COVID-19 case compared to White decreased for all demographics in interval 3: the risk of cases in Black demographics was equal to or lower than White, but risk remained elevated for all Hispanic/Latinx demographics (Figure 4D).

### Temporal trends in county-level COVID-19 case disparities by age

We assessed relative risk of a COVID-19 case in non-White vs White individuals at the county level for each age group and each of the 3 epidemic intervals (Figure 5). When significant, relative risk was always higher in the non-White than in the White population.

**Figure 5.**
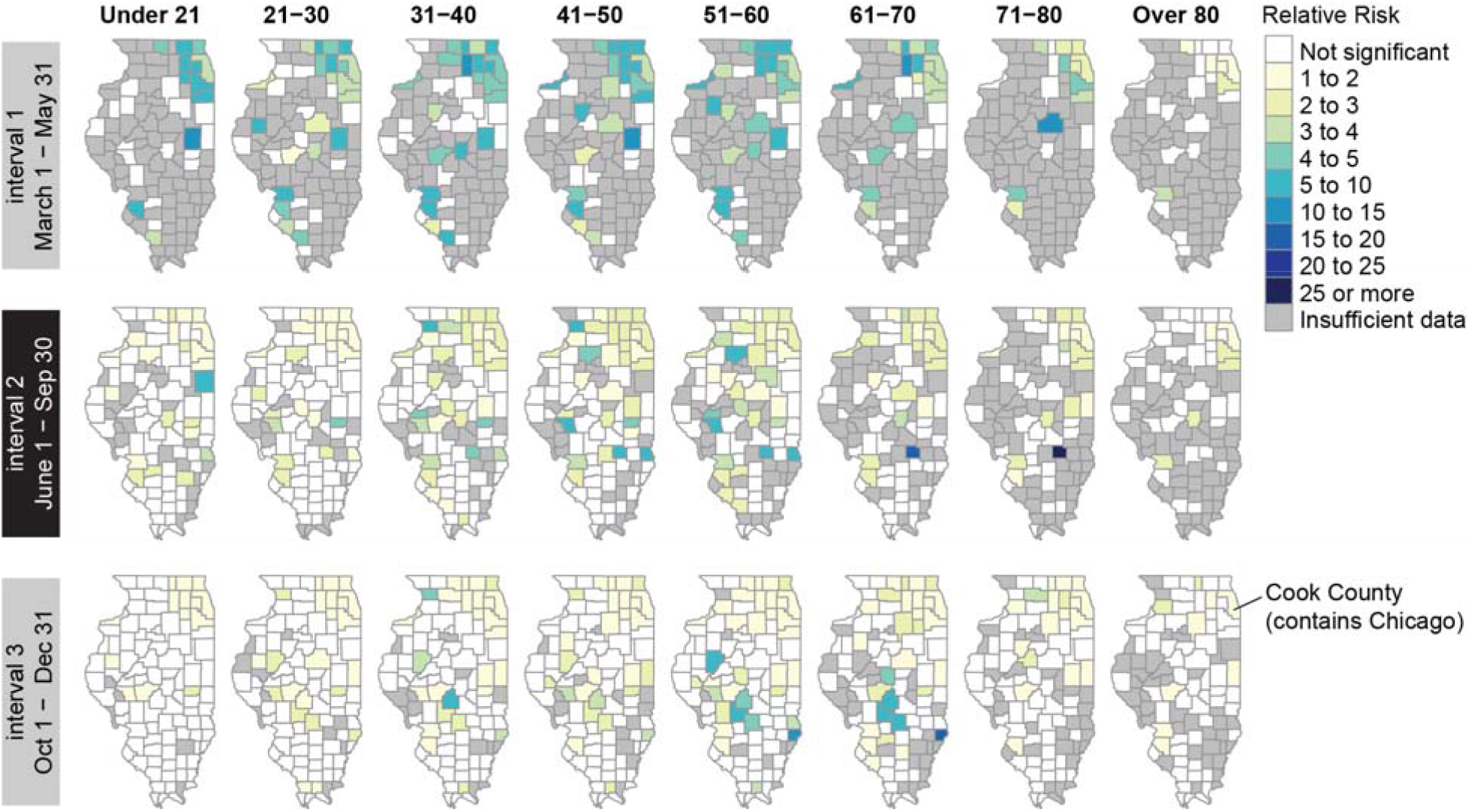
Relative risk of COVID-19 cases for non-White individuals compared with White individuals, by county and age group, during each epidemic interval from March 1 to December 31, 2020. Color indicates x-fold increase in risk for non-White vs. White residents. Counties colored white had relative risk that was not significantly different from 1 based on a 95% CI. Relative risk was not calculated (gray) when a minimum of 10 non-White cases and 10 White cases was not met.

Cases in interval 1 were primarily located in northeastern Illinois around Cook County, where Chicago is located, and in southwestern counties across the border from St. Louis, Missouri. Disparities were highest in the counties surrounding Cook County and in the southwest. During intervals 2 and 3, the epidemic spread throughout the state, and the magnitude of disparities decreased across age groups in most counties impacted during interval 1. However, relative risk remained above 1 in the northeast counties for all age groups. In interval 3, disparities were highest among working-age adults in several counties in south central Illinois.

### Temporal trends in age-adjusted disparities in risk of a COVID-19 death

Black and Hispanic/Latinx people in Illinois died from COVID-19 at higher rates than White people in 2020 (Figure 6). Losses were particularly high among minority elders: cumulatively 0.88% and 1.04% of Illinois’ Black and Hispanic/Latinx populations over the age of 60 died from COVID-19 in 2020, compared to 0.54% of the White population over the age of 60.

**Figure 6.**
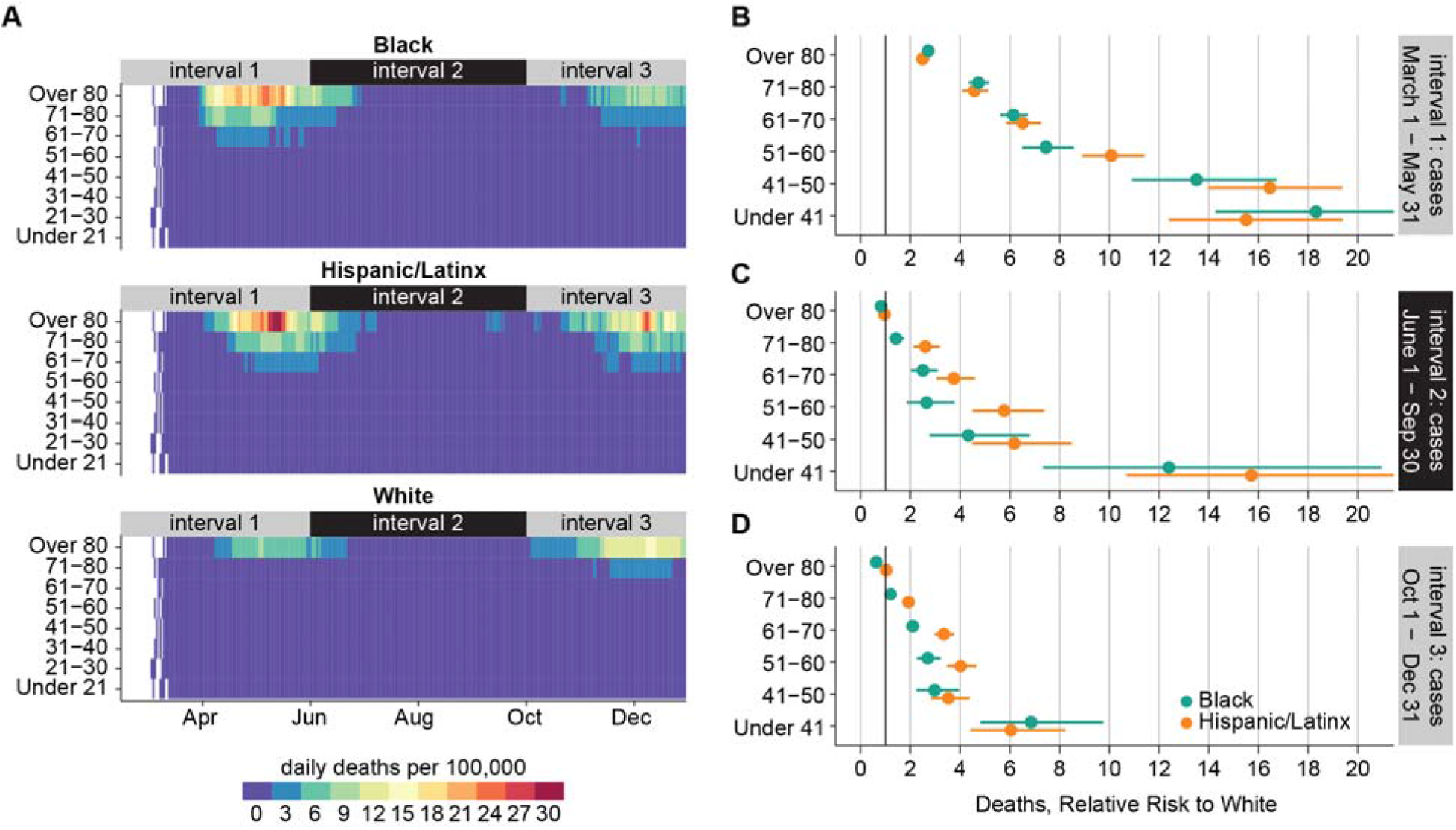
Age-adjusted disparities in risk of COVID-19 death by race/ethnicity from March 1 to December 31, 2020. A) 7-day moving average of COVID-19 deaths per 100,000 population, by age and race/ethnicity. B-D) Relative risk of COVID-19 death over three epidemic intervals. Horizontal lines indicate 95% confidence intervals. Deaths are binned into intervals based on the date of case, and interval 3 is truncated to remove lagged deaths.

Disparities in COVID-19 mortality were largest during interval 1. Throughout the epidemic, the daily per capita death rate remained below 3 per 100,000 for White people under 71. In interval 1, death rates exceeded 3 per 100,000 for Hispanic/Latinx and Black people ages 61-70, 9 per 100,000 for those ages 71-80, and 24 per 100,000 for those over 80. For one week in May, the death rate for Hispanic/Latinx people over 80 surpassed 30 per 100,000.

COVID-19 mortality rates during interval 1 were greater in all Hispanic/Latinx and Black demographics than in White, and the relative risk of death increased with younger age (Figure 6B). Relative risk of COVID-19 death in the Hispanic/Latinx population was significantly higher than in the Black population for the 51-60 age group but not significantly different in other groups. While the absolute risk of death was low for younger adults, Hispanic/Latinx and Black populations under 41 had mortality rates more than 12 times that of the White population during interval 1. Risk of COVID-19 death for Black and Hispanic/Latinx compared to White was 4-6 times higher for those 61-80 and 2 times higher for those over 80.

In interval 2, daily per capita death rates fell below 3 per 100,000 for all demographics except Hispanic/Latinx people over 80. Death rates began to rise again in early October during interval 3. The daily death rate among Hispanic/Latinx people over 80 reached 27 per 100,000 in early December, and death rates were also higher for Hispanic/Latinx individuals 51-80 compared to Black and White populations. In December 2020, the daily death rate for the White over-80 population rose above 15 per 100,000: the highest rate seen in this demographic in 2020. Death rates for Black people of all ages remained below 12 per 100,000 during interval 3.

The relative risk of COVID-19 death for Black and Hispanic/Latinx people compared to Whites fell during intervals 2 and 3 (Figure 6C, 6D), reaching 1 for the over-80 age group. Disparities persisted for ages below 70. At the end of 2020, relative risk of COVID-19 death compared to White for ages 51-80 remained significantly higher for Hispanic/Latinx individuals than for Black individuals the same age. Risk of COVID-19 death in individuals under 41 was still 5-10 times higher for Black and Hispanic/Latinx individuals than for White.

To investigate the drivers of disparities in COVID-19 mortality rate, we considered counterfactuals where minoritized populations experienced incidence, case fatality ratio (CFR), or mortality rate at the same level as White people (Figure 7A). All three counterfactuals reduced mortality for minoritized populations during interval 1, and counterfactual incidence reduced mortality more than did counterfactual CFR. This difference suggests that the disparities in COVID-19 mortality early in the epidemic were driven primarily by higher exposure of minoritized populations to SARS-CoV-2, rather than elevated risk of death given detected infection.

**Figure 7.**
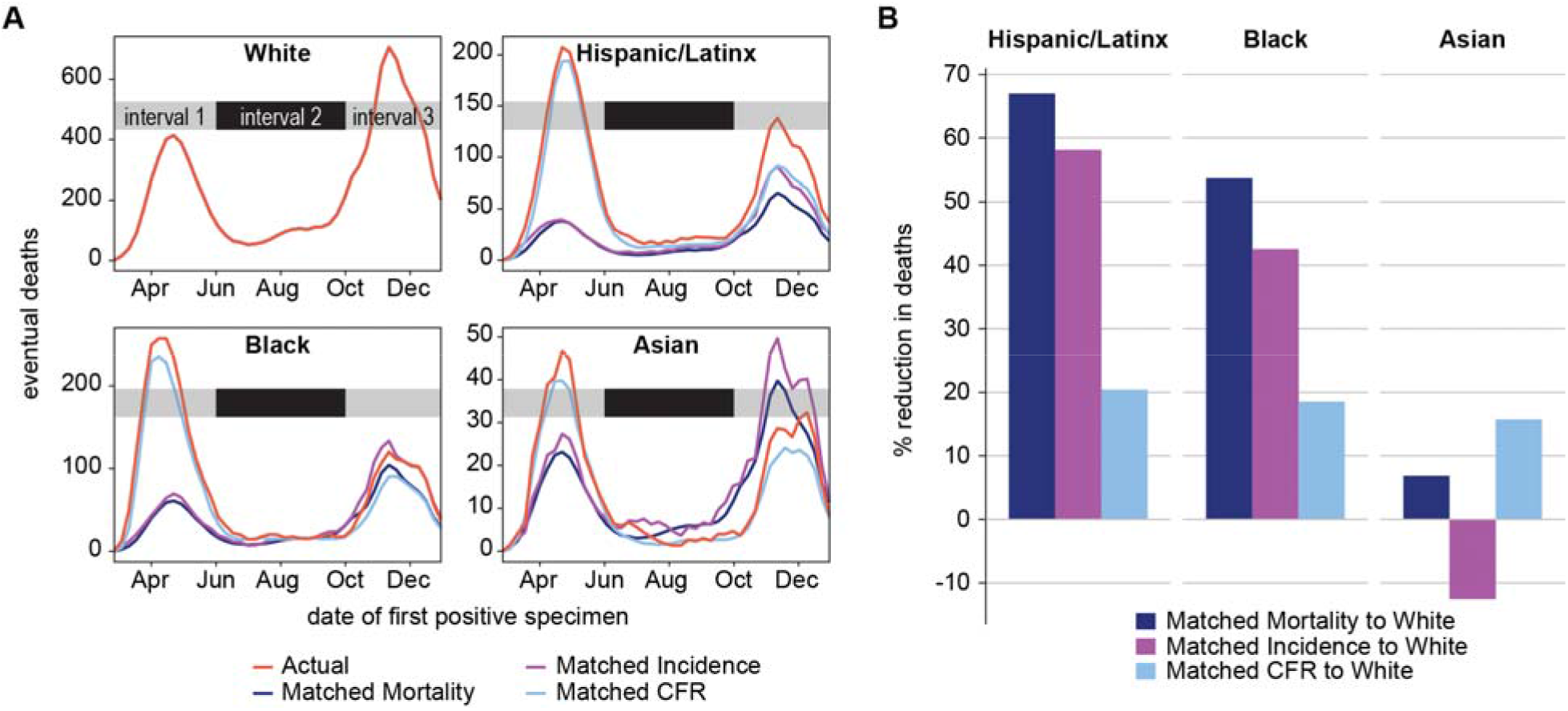
COVID-19 deaths attributable to differences in age-adjusted incidence, case fatality ratio, or both, between Asian, Black, and Hispanic/Latinx populations compared to White. A) Weekly actual and counterfactual deaths by date of first positive specimen. B) Cumulative percent reduction of deaths, counterfactual vs. actual deaths.

Starting in June for Asian and September for Black, counterfactual deaths predicted using White incidence begin to exceed the actual deaths observed, reflecting an increased incidence in White populations that outpaced incidence in Asian and Black demographics. Counterfactual deaths remained less than actual deaths among Hispanic/Latinx people, regardless of parameter substitution, indicating continued elevated exposure risk in this demographic.

Cumulatively, substituting age-matched White incidence without adjusting CFR reduced Black deaths by 1,424 (42.5%) and Hispanic/Latinx deaths by 1,711 (58.1%) (Figure 7B). Substituting White CFR without adjusting incidence only reduced deaths in these groups by 18-20%. For Asians, substituting White case fatality ratio resulted in fewer deaths than have been reported, although substituting White incidence had the opposite effect. Had they experienced the same average per capita mortality rate as White demographics each week, 1,972 Hispanic/Latinx people, 1,796 Black people, 44 Asian people, 16 Native people, and 79 people of “Other” race would not have died of COVID-19 (Figure 7). By this approach, racial/ethnic disparities account for 3,907 total lives lost to COVID-19 in Illinois in 2020.

## Discussion

Minoritized groups experienced an outsized burden of COVID-19 morbidity and mortality in Illinois during 2020, primarily due to differences in exposure risk. Black and Hispanic/Latinx cases were distributed across age groups or concentrated in working age adults throughout 2020, while White cases were concentrated in the elderly in early 2020 and in young adults in mid-2020. Although racial disparities in Illinois lessened following the initial epidemic interval from March through May 2020, Hispanic/Latinx and Black individuals remained significantly more likely to die of COVID-19 through the end of the year in all but the oldest age groups. Given the unknown bias in surveillance quality with respect to cases and TPR as well as the lack of robust hospitalization data, confirmed COVID-19 deaths are the most robust primary outcome to capture the evolution of racial/ethnic disparities in Illinois in the period being studied.

If incidence becomes more equitable, structural racism can still cause differences in case fatality and contribute to more deaths among minoritized populations. One consequence of residential segregation is variable access to high-quality care, preventive interventions, and accurate information [18]. This combined with racial-occupational disparities in health insurance and worker’s compensation claim payouts [19] could lead to delays in care and worse outcomes. It is also possible that extant disparities in certain comorbid conditions like diabetes and hypertension [20]-[21] or biased treatment [22] contribute to poorer prognosis for hospitalized Black and Hispanic/Latinx COVID-19 patients. The convergence of COVID-19 incidence and mortality rate in the elderly population for all three racial and ethnic groups in late 2020 suggests that infection fatality rates are likely to be similar across the groups and that prevalence of comorbidities may play only a minor role in explaining disparities in COVID-19 deaths. Our finding that disparities in COVID-19 burden were primarily driven by differences in exposure rather than differences in case fatality rate, similar to what was found in Michigan in early 2020 [23], underscores the importance of providing access to and support for COVID-19 prevention in minoritized communities.

The low quality of data on hospitalizations in I-NEDSS did not permit analysis of racial and ethnic disparities in incidence of severe COVID-19 infection. Therefore, we could not distinguish between various drivers of case fatality such as access to care, quality of care, and disease severity. An analysis of COVID-19 outcomes from a health system in California, Oregon, and Washington found increased odds of testing positive and hospitalization for all minoritized races/ethnicities, though only Hispanic/Latinx ethnicity was associated with in-hospital mortality, attributed to delays in accessing care [24].

We have presented evidence of racial and ethnic disparities in diagnostic testing rates, detected cases, hospital admissions, and confirmed deaths, but our ability to accurately capture the many possible contributions of structural racism to COVID-19 disparities is limited by the data available. Case detection rates were biased by testing patterns and symptom severity, especially early in the epidemic when access to diagnostic testing was very limited. Reason for testing was not systematically reported, obfuscating the interpretation of testing rates and TPR between epidemic intervals and demographic groups with different rates of routine (asymptomatic) testing. In 2020, routine diagnostic testing was conducted in congregate settings such as university communities and among long-term care facility residents and staff. It is possible that different demographic groups were subject to routine testing at different rates and thus routine testing would differentially affect demographic-specific TPR and cases. However, we show that high relative risk of COVID-19 cases among working-age Hispanic/Latinx and Black adults from March to May 2020 was not due to a higher case detection rate in these populations. At the end of 2020, Hispanic/Latinx people of all ages in Illinois remained over-burdened by, and under-tested for, COVID-19 relative to non-Hispanic Whites.

Per capita case and death rates were calculated using estimates from the American Community Survey, which includes only the civilian non-institutionalized population. State and county-level disparities may be skewed by cases among incarcerated/detained people, residents of developmental and long-term care facilities, undocumented immigrants, and active-duty military members as these groups are excluded from ACS population estimates.

Demographic information was provided to I-NEDSS from various mandated reporters and was not reliably collected in the same way for all patients. Ethnicity was missing for nearly 40% of cases, raising concern for under-counting of cases and deaths as Hispanic/Latinx in this dataset. 43.9% of tests, 29.2% of cases, and 3.5% of deaths were missing both race and ethnicity. Cases with and without race/ethnicity did not differ significantly in their age distributions, nor in the majority race of their home ZIP code populations, suggesting that the missing race/ethnicity data may not be systematically biased at the state level (Supp Figure 2), suggesting that the analyses shown in this study are representative of true trends across Illinois. Analyses presented in this paper assumed non-Hispanic/Latinx ethnicity for all cases of unknown ethnicity. A sensitivity analysis (Figure S3) found that this assumption leads to more conservative relative risk estimates – exclusion of cases with unknown ethnicity gives higher relative risk estimates than those presented.

As this pandemic progressed, two major state and local government-led responses called for embedding equity into COVID-19: the IDPH COVID-19 Equity Team and the Chicago Mayor’s Racial Equity Rapid Response Team. These initiatives were data-driven, community partnered teams assembled by the Illinois Governor, IDPH, and the Chicago Mayor to address and mitigate COVID-19 illness and death in Black and Brown communities across Illinois. The overarching goals of both initiatives were not only to reduce infection and mortality COVID-19 disparities but also to build the groundwork for future work to address the longstanding systemic inequities in Illinois’ communities of color.

Race and ethnicity are socially constructed, and the demographic stratifications employed in this study are commonly used categories that do not necessarily reflect shared experiences or circumstances. Race itself is not a biological risk factor: rather, racial health disparities are the result of generations of structural racism and barriers to health care access, and many possible drivers of disparities have been proposed. Evaluating the extent to which exposure, rather than case fatality ratio, has driven disparities in COVID-19 requires a systematic exploration of the mechanisms by which structural racism drives the behavioral, economic, social, and health factors that kill minoritized populations during a global pandemic. Describing trends in COVID-19 disparities thus far can help identify populations to monitor for long-term implications of disparate COVID-19 burden and inform future epidemic responses that center underserved and over-burdened Black and Hispanic/Latinx communities.

Future studies can build upon this work by probing the specific structural drivers of racial/ethnic disparities at high spatial resolution, such as neighborhood-level social vulnerability or environmental justice indices. Our analyses include only cases detected through the end of 2020, before vaccines that protect against COVID-19 were widely available. A similar investigation of disparities in later months is needed to understand how vaccination uptake affected the epidemic in different communities.

## Conclusions

During the first epidemic wave in March to May 2020, Black and Hispanic/Latinx populations were 2.5-5 times and 2.5-8.5 times more likely to experience a COVID-19 case, respectively, and 2-18 times more likely to die of COVID-19, compared to age-matched White populations. Racial and ethnic disparities in COVID-19 deaths were driven primarily by higher risk of exposure for minoritized populations, which can be explained by underlying structural racism.

## Data Availability

The I-NEDSS and testing by age and race/ethnicity datasets analyzed in this study were used under license for the current study, and so are not publicly available. Restrictions apply to the availability of these data, which contain identifiable private health information. Interested parties should contact IDPH to inquire about access. Public data on cases and testing are available from IDPH.

https://www.dph.illinois.gov/covid19/covid19-statistics

## List of abbreviations

ACS: American Community Survey
CFR: case fatality ratio
CLI: COVID-like illness
IDPH: Illinois Department of Public Health
I-NEDSS: Illinois’s National Electronic Disease Surveillance System
TPR: test positivity rate

## Declarations

### Ethics approval and consent to participate

This study was carried out as part of a Medical Study “Modeling COVID-19 Epidemiologic Trend and Health Care Impact in Illinois” declared by IDPH on March 23, 2020. All data collection was performed by IDPH as part of routine surveillance for COVID-19 and was deidentified prior to analysis. The Northwestern University Institutional Review Board has ruled that this study does not constitute human subjects research.

### Consent for publication

Not applicable

### Availability of data and materials

The I-NEDSS and testing by age and race/ethnicity datasets analyzed in this study were used under license for the current study, and so are not publicly available. Restrictions apply to the availability of these data, which contain identifiable private health information. Interested parties should contact IDPH to inquire about access. Public data on cases and testing are available from IDPH (https://www.dph.illinois.gov/covid19/covid19-statistics).

### Authors’ contributions

TMH and JG conceived the project. TMH completed all analyses. TMH and JG wrote the initial draft. All authors revised and approved the final manuscript.

## Acknowledgements

We thank Stacey Hoferka Jensen, Dejan Jovanov, and Sara Rogers for data extraction and preparation from I-NEDSS, and Kiarri Kershaw and Mercedes Carnethon for helpful discussions. We thank Kemba Noel-London for helpful comments on the manuscript.

