## Supplemental Figures for "Structural racism and COVID-19 response: Higher risk of exposure drives disparate COVID-19 deaths among Black and Hispanic/Latinx residents of Illinois, USA"

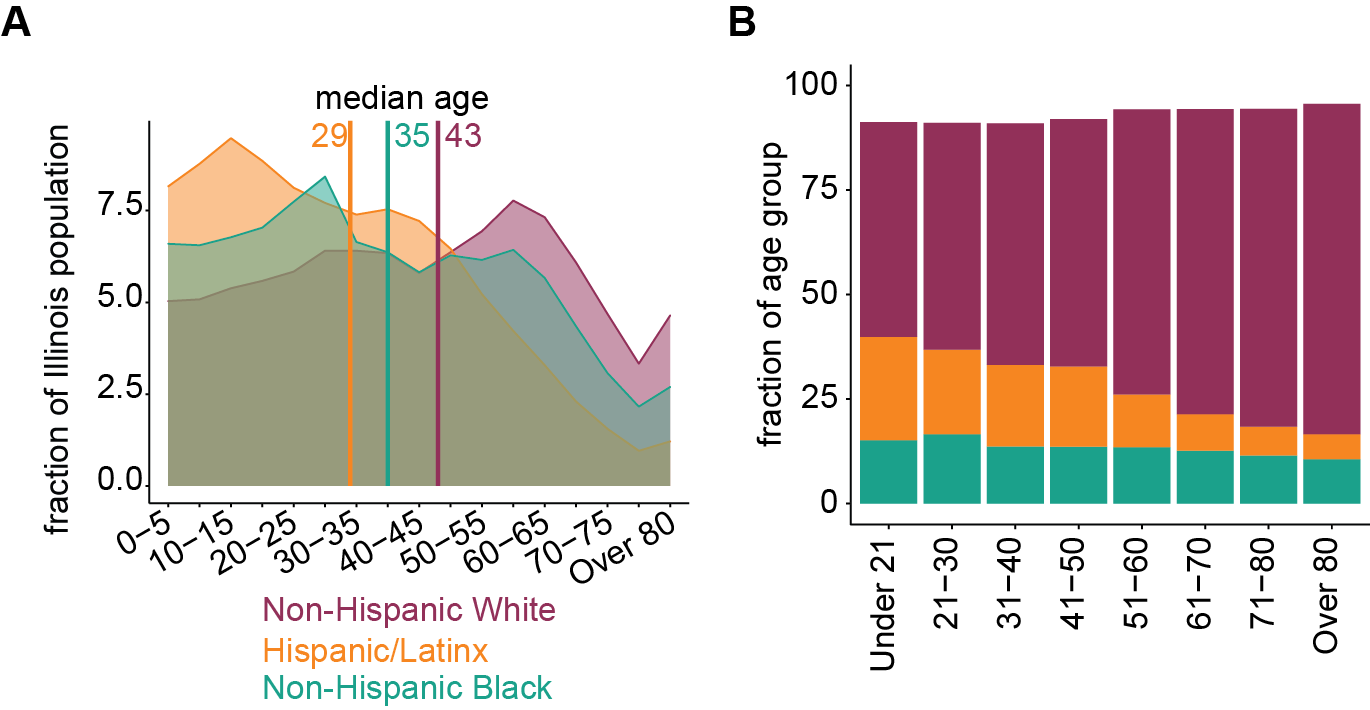

Figure S1. Demographics of Illinois by age and race. A) Age distribution by race/ethnicity. B) Racial/ethnic distribution by age.

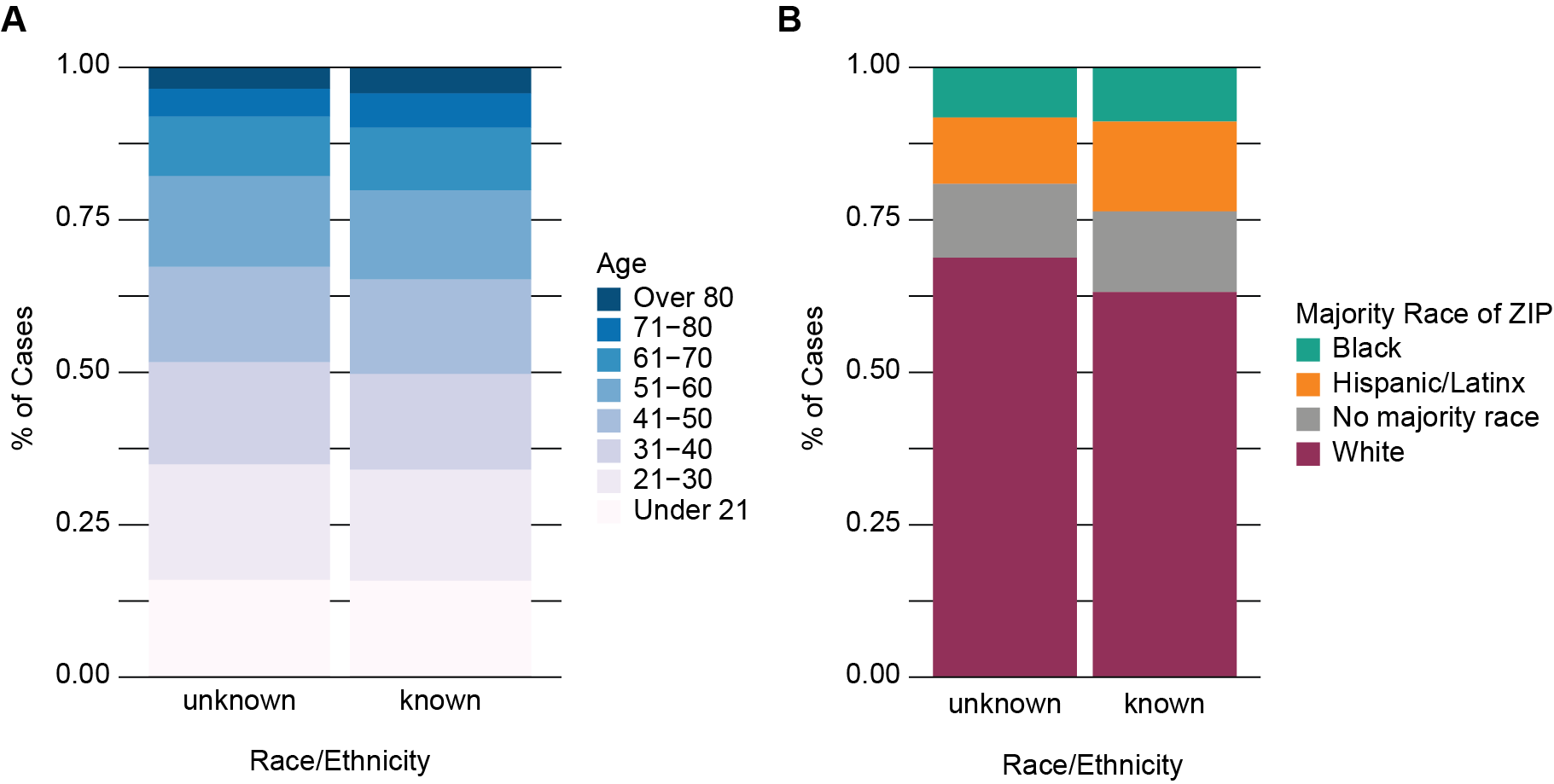

Figure S2. Demographics of COVID-19 cases with vs. without reported race/ethnicity. A) Age distribution. B) Majority race of ZIP code.

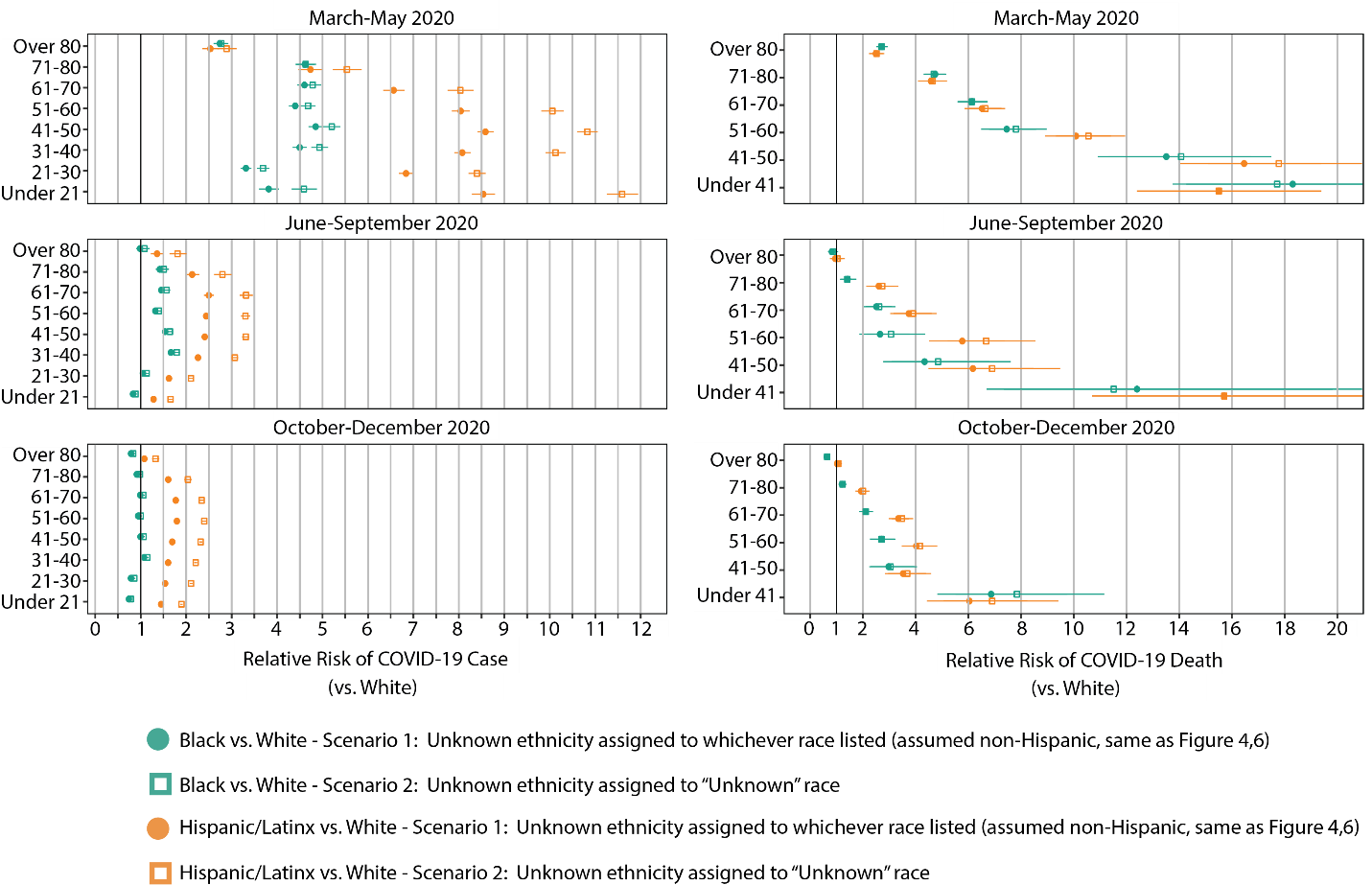

Figure S3. Sensitivity of Relative Risk of A) COVID-19 Cases and B) COVID-19 Deaths to case race/ethnicity assignment Scenarios were run in which cases with “unknown” ethnicity were: 1) Assumed to be non-Hispanic and assigned to the recorded racial group (solid circle, and identical to Figures 4B-D and 6B-D), or 2) Assigned to the “unknown” racial group (open square. In most cases, the default assumption of non-Hispanic ethnicity made in main text above results in a lower relative risk of COVID-19 deaths and significantly lower relative risk of COVID-19 cases for Black and Hispanic/Latinx populations, than if cases of unknown ethnicity were excluded.
